# Adverse childhood experiences and multiple mental health outcomes through adulthood: a prospective birth cohort study

**DOI:** 10.1101/2021.02.23.21252273

**Authors:** Dawid Gondek, Praveetha Patalay, Rebecca E Lacey

**Affiliations:** Research Department of Epidemiology and Public Health, University College London, London, England; Centre for Longitudinal Studies, Department of Social Science, UCL Institute of Education, University College London, London, England; MRC Unit for Lifelong Health and Ageing, Department of Population Science and Experimental Medicine, University College London, London, England

**Keywords:** adverse childhood experiences, mental health, psychological distress, wellbeing, life course, National Child Development Study

## Abstract

This study examined the association between adverse childhood experiences (ACEs) and mental health-related outcomes spanning ages 16-55 years, using a prospective birth cohort representative of those born around 1958 in Great Britain.

We found a dose-response association between prospectively and retrospectively reported ACEs and all studied mental health-related outcomes, after accounting for multiple covariates. Among those with 2+ (vs 0) prospective ACEs, the risk of clinically significant psychological distress was up to 2.14 times higher, and of seeing a mental health specialist up to 2.85 times higher. Our findings reiterate the need for early-life interventions to reduce inequalities in mental health.

## Introduction

Exposure to adverse childhood experiences (ACEs) is associated with a broad range of mental health-related outcomes.^1-4^ However, previous studies tended to use retrospectively reported ACEs, potentially suffering from recall bias, and measured mental health outcomes at only one-time point in adulthood, precluding understanding of the persistence of any impacts of ACEs on mental health across adulthood.^1^ Besides, existing longitudinal research on ACEs is focussed exclusively on mental ill-health in the form of general psychological distress.^1,2^ While the impact of ACEs on other aspects of mental health throughout adulthood is less known, for instance, wellbeing, utilisation of mental health services, or taking psychotropic medications.^2^

These limitations can be addressed by taking advantage of rich longitudinal data available in the 1958 National Child Development Study (NCDS),^5^ a prospective birth cohort representative of those born around 1958 in Great Britain. The main objective of our study was to examine the association between ACEs, reported both prospectively and retrospectively, and a broad range of mental health-related outcomes spanning age 16-55 years while accounting for multiple covariates. The outcomes included utilisation of mental health services, taking psychotropic medications, standardised measures of psychological distress, life satisfaction, quality of life, and self-reported general health (as it tends to correlate strongly with mental health).^6^ Inclusion of these outcomes also allowed for direct comparison of the relative magnitude of the association between ACEs and various domains of mental health, which to our knowledge has not been studied previously.

## Methods

We limited the sample of the NCDS^5^ to those without missing information on retrospectively reported ACEs and based in Great Britain at the last data wave (at age 55) (n=7,980) (see eFigure 1 for sample flow diagram).^5^ We included ACEs identified by previous studies using NCDS,^7,8^ and considered them cumulatively as an ACE score. The prospective ACE score ranged from ‘0 ACEs’ to ‘2+ ACEs’, and retrospective from ‘0 ACEs’ to ‘4+ ACEs’. See Fig. 1 for the list of individual ACEs and eTable 1 for more details on how they were measured.

**Figure 1.**
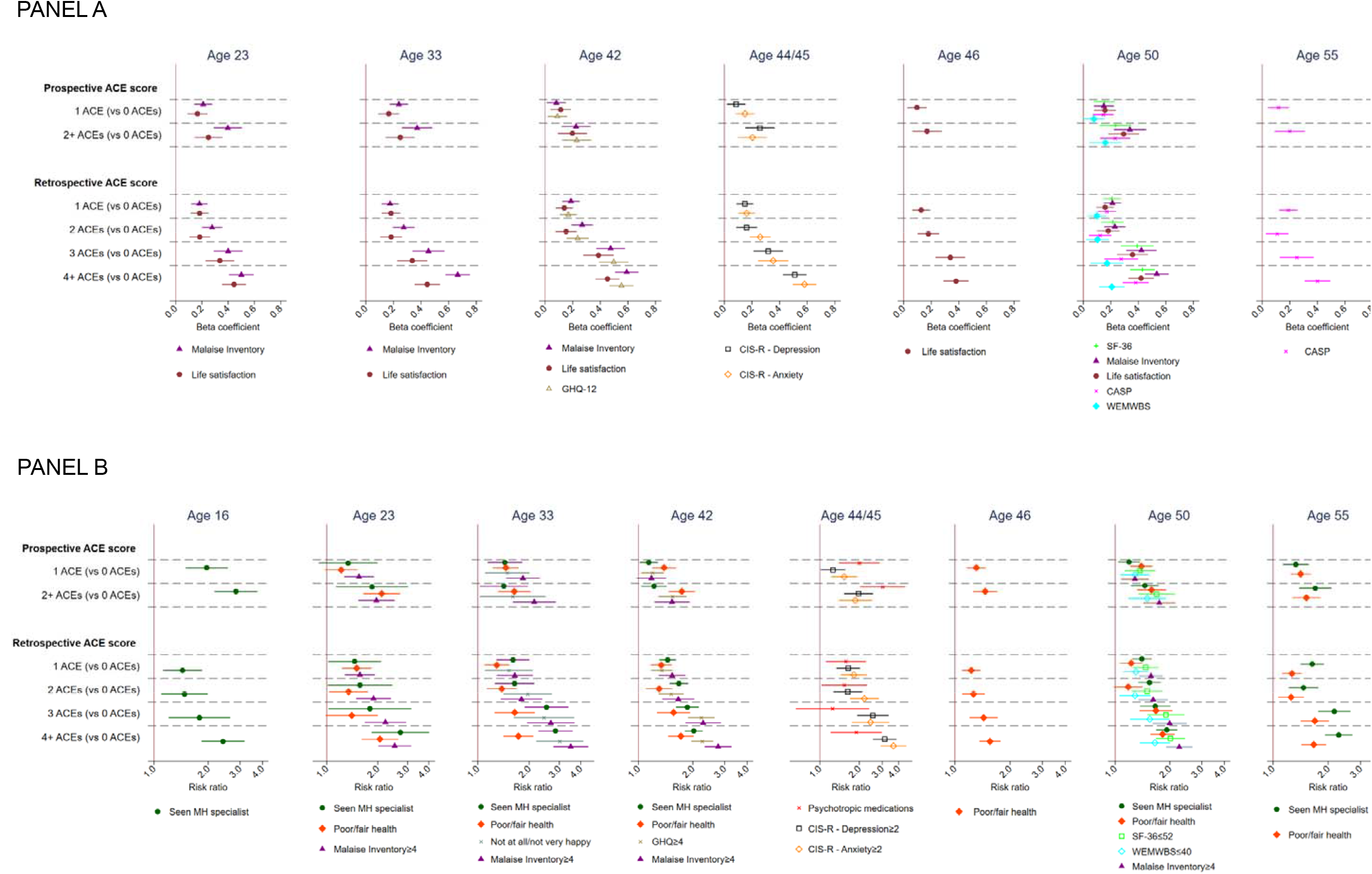
The association between the ACE score and mental health – continuous (Panel A), binary (Panel B) outcomes. *Note*. Prospectively measured ACEs included physical neglect (reported by teacher at age 7, 11), parental separation/divorce, parental substance misuse, family conflict, death of parent, parental mental health problems, parental offending (all reported by health visitor and/or parent at age 7-16). Retrospectively measured ACEs were parental separation/divorce (reported by cohort member at age 33), parental substance misuse, family conflict, witnessing abuse, parental mental health problems, sexual abuse, physical abuse, psychological abuse, emotional neglect (all reported by cohort member at age 44/45). The estimates were adjusted for covariates – gender, father’s occupational social class, maternal education, birthweight, gestational age, maternal age at birth and breastfeeding duration.

Continuous measures of psychological distress included the Malaise Inventory (at age 23, 33, 42, 50), General Health Questionnaire (GHQ-12; at age 42), depression and anxiety subscales of Clinical Interview Schedule - Revised (CIS-R; at age 44/45), psychological distress and well-being subscale of the Short Form Health Survey (SF- 36; at age 50). Continuous measures of wellbeing comprised the Warwick-Edinburgh Mental Wellbeing Scale (WEMWBS; at age 50), and Quality of Life Scale (CASP; 12- item at age 50; 6-item at age 55).^9^ All measures were standardised into Z scores and wellbeing-oriented scales were reversed in line with higher scores on psychological distress measures indicating worse mental health.

Binary outcomes included seeing a mental health specialist (at age 16, 23, 33, 42, 50 55), poor/fair (vs good/very good/excellent) self-reported general health (at age 23, 33, 42, 46, 50 55), being not at all/not very happy (vs fairly/very happy) (at age 33), indicators of clinically significant psychological distress – the Malaise Inventory (score≥4), GHQ (score≥4), CIS-R depression/anxiety (score≥2), SF-36 (score≤52), and WEMWBS (score≤40).^9^ See eTable 2 for details on all outcome measures.

The associations between the ACE score and mental health were studied with covariates-adjusted (see Figure 1 for the list of covariates) linear regression for continuous and robust Poisson regression for binary outcomes, based on the multiple imputation with chained equations (20 datasets).^10^ Men and women were combined, as we found no evidence for differential ACEs-mental health associations between genders. The NCDS has been granted ethical approval for each sweep from 2000 by the National Health Service (NHS) Research Ethics Committee. All participants provided written informed consent after a thorough explanation of the research procedures.

## Results

We found strong evidence for a dose-response association between prospectively and retrospectively reported ACEs and all mental health-related outcomes, after accounting for multiple covariates (Fig. 1). The associations were robust to additional adjustment for internalising and externalising symptoms at age 7 (results not shown) and their magnitude was relatively consistent regardless of ACEs being reported prospectively or retrospectively. For brevity, findings on the prospective ACE score are discussed (see Fig. 1 for all results).

< Figure 1. The association between the ACE score and mental health – binary (Panel A), continuous (Panel B) outcomes.>

The prospective ACE score of 2+ (vs 0) was associated with between 0.160 (95% CI 0.044 to 0.276) and 0.293 (95% CI 0.182 to 0.405) standard deviations (SD) higher score on measures of wellbeing, including life satisfaction, CASP, and WEMWBS. Scores on the measures of psychological distress, including the Malaise Inventory, CIS-R, GHQ-12, and SF-36, were between 0.203 (95% CI 0.099 to 0.308) and 0.398 (95% CI 0.290 to 0.505) SD higher. ACE score tended to have a somewhat stronger association with psychological distress than wellbeing when measured at the same age. For instance, at age 50 a retrospective ACE score of 4+ (vs 0) was associated with 0.534 (95% CI 0.448 to 0.621) SD higher score on the Malaise Inventory, compared with 0.208 (95% CI 0.114 to 0.301) on WEMWBS. The correlation of measures within the same domains of mental health was moderate when collected at the same age (r=0.56-0.68 for psychological distress and r=0.49-0.69 for wellbeing) (see eTable 3 for all correlations). The correlations between wellbeing and distress measures tended to be weak (r=0.16-0.54) when collected at the same age, with SF- 36 having a moderate correlation with both wellbeing and distress measures (r=0.52- 71).

The prospective ACE score of 2+ (vs 0) was associated with between 50% (risk ratio, 1.50, CI 95% 1.19 to 1.90) and 2.14 times (CI 95% 1.60 to 2.86) higher risk of having clinically significant psychological distress and up to 2.85 times (CI 95% 2.17 to 3.75) higher risk of seeing a mental health specialist. The correlation between measures of distress and seeing a mental health specialist was moderate (rho=0.45-0.63) when collected at the same age. The correlation between measures indicating clinically significant distress was moderate to strong (rho=0.62-0.80).

Those with a prospective ACE score of 2+ (vs 0) had also 3 times higher risk of taking psychotropic medications at age 44/45 (CI 95% 2.03 to 4.50). Taking medications had a weak correlation with CIS-R depression (r=0.17) or anxiety (r=0.16). The prospective ACE score of 2+ (vs 0) was associated with 60% (CI 95% 1.03 to 2.50) higher risk of not being happy at all or not very happy (vs fairly/very happy) at age 33 and with up to 2.11 times (95% CI 1.64 to 2.71 at age 23) higher risk of poor or fair (vs good/very good/excellent) self-rated general health. Having poor or fair self-rated general health had a weak to moderate association with clinically significant psychological distress (rho=0.35-0.53) and seeing a mental health specialist (rho=0.32-0.56).

## Discussion

We found that being exposed to ACEs is associated, in a dose-response fashion, with a range of mental health-related outcomes between age 16 and 55, thus replicating and extending previous research.^1-3^ These associations appear to be somewhat stronger for psychological distress than wellbeing, are robust to rich covariates-adjustment, including early-life emotional development, and are not sensitive to ACEs being measured prospectively or retrospectively, or outcomes being collected at specific ages. The effect sizes in our study for those with 4+ retrospective ACEs were comparable to those obtained by the meta-analysis by Hughes and colleagues, for self-rated health, anxiety, depression, and life satisfaction.^1^

The main strength of our study was that is used measures of various domains of mental health spanning age 16-55 and capturing self-reported psychological distress, wellbeing and impact of ACEs on the utilisation of mental health services, however, it is a limitation that not all measures under investigation were available at each age. To mitigate any potential bias due to non-response or attrition, we multiply imputed missing values while including a rich set of predictors of missing information in the imputation model. This allowed for imputing missing data with greater precision.^11^

Our findings reiterate the need to address early-life adverse events to reduce inequalities in a broad range of adult mental health outcomes across adulthood.

## Data Availability

Data are available from UK Data Services

## Article Information

### Author Contributions

Dawid Gondek had full access to all of the data in the study and takes responsibility for the integrity of the data and the accuracy of the data analysis.

### Study concept and design

Dawid Gondek, Praveetha Patalay, Rebecca Lacey. Acquisition, analysis, or interpretation of data: Dawid Gondek, Praveetha Patalay, Rebecca Lacey.

Drafting of the manuscript: Dawid Gondek.

Critical revision of the manuscript for important intellectual content: Dawid Gondek, Praveetha Patalay, Rebecca Lacey.

### Statistical analysis

Dawid Gondek.

Administrative, technical, or material support: Dawid Gondek, Praveetha Patalay, Rebecca Lacey.

### Study supervision

Dawid Gondek, Praveetha Patalay, Rebecca Lacey.

