## Supplemental material for "Adverse childhood experiences and multiple mental health outcomes through adulthood: a prospective birth cohort study"

**Supplementary Material**

**Did not complete retrospective ACE questionnaire (at age 44/45):** n = 9,269

**Original sample**

**(born in Great Britain):**

n = 17,415

n = 8,146

**Died or emigrated by the last data sweep (age 55):** n = 166

**Analytical sample:**

n = 7,980

eFigure 1. Sample flow diagram

| eTable 1. Measures of prospectively and retrospectively reported adverse childhood experiences (ACEs). | | | |
| --- | --- | --- | --- |
| **Adversity** | **Age collected** | **Informant** | **Description** |
| **Prospectively reported ACEs** | | | |
| Parental separation/divorce | 7 | Health visitor | Divorce or separation listed as difficulty of the family |
|  | 11, 16 | Parent | Relationship to child of person acting as child's parents & reason for change |
| Parental substance misuse | 7 | Health visitor | Alcoholism listed as difficulty of the family |
| Family conflict | 7 | Health visitor | Domestic tension listed as difficulty of the family |
| Death of parent | 7 | Health visitor | Death of mother listed as difficulty of the family |
|  | 7 | Health visitor | Death of child's father listed as difficulty of the family |
|  | 11, 16 | Parent | Relationship to child of person acting as child's parents & reason for change |
| Parental mental health problems | 7 | Health visitor | Mental illness or neurosis listed as difficulty of the family |
|  | 7 | Health visitor | Family uses services of a psychiatric social worker |
|  | 7, 11, 16 | Parent | Mother or father has a chronic mental illness |
| Physical neglect | 7, 11 | Teacher | Child appears scruffy/dirty/underfed |
| Parental offending | 7 | Health visitor | Family in contact with probation services |
|  | 11 | Parent | Family in contact with probation services or family member in prison |
|  | 16 | Parent | Family member in contact with probation services |
| **Retrospectively reported ACEs** | | | |
| Parental separation/divorce | 33 | Cohort member | Parents ever permanently separated or divorced & how old when happened |
| Parental substance misuse | 44/45 | Cohort member | Mother had trouble with drinking or other drug use |
|  | 44/45 | Cohort member | Father had trouble with drinking or other drug use |
| Family conflict | 44/45 | Cohort member | There was much conflict and tension in the household whilst I was growing up |
| Witnessing abuse | 44/45 | Cohort member | I witnessed physical or sexual abuse of others in my family |
| Parental mental health problems | 44/45 | Cohort member | Mother or father suffered from nervous or emotional trouble or depression |
| Sexual abuse | 44/45 | Cohort member | I was sexually abused by a parent |
| Physical abuse | 44/45 | Cohort member | I was physically abused by a parent - punched, kicked or hit or beaten with an objective, or needed medical treatment |
| Psychological abuse | 44/45 | Cohort member | I was verbally abused by a parent |
|  | 44/45 | Cohort member | I suffered humiliation, ridicule, bullying or mental cruelty from a parent |
| Emotional neglect | 44/45 | Cohort member | My mother was unaffectionate |
|  | 44/45 | Cohort member | My father was unaffectionate |

eTable 2. Details on measures of mental health-related outcomes.

Malaise Inventory (9-item)^1^ (age 23, 33, 42, 50); ranging 0-9. Clinical case at score ≥4.

Cohort member has seen a specialist for mental health problems since previous data sweep (age 16, 23, 33, 42, 50, 55)

Poor/fair (vs good/very good/excellent) self-rated general health (age 23, 33, 42, 46, 50, 55)

Life satisfaction* (age 23, 33, 42, 46, 50)

On a scale of 0 ("Completely dissatisfied") to 10 ("completely satisfied") with the way life has turned out so far.

At age 33, the question was retrospective - about life satisfaction at age 23.

General Health Questionnaire (GHQ-12)^2,3^ (age 42); ranging 0-12. Clinical case at score ≥4.

Depression and anxiety subscales of Clinical Interview Schedule - Revised (CIS-R)^4^ (age 44/45); ranging 0-4. Clinical case at score ≥2.

General mental health (psychological distress and well-being) of the Short Form Health Survey (36-item) (SF-36)*^5^ (age 50); ranging 0-100. Clinical case at score ≤52.

Warwick-Edinburgh Mental Wellbeing Scale (WEMWBS)*^6,7^ (age 50); ranging 0-70. Clinical case at score ≤40.

Quality of Life Scale (CASP; 12- item at age 50; 6-item at age 55)*^8,9^; ranging 2-36 at age 50 and 6-24 at age 55.

Psychotropic medications self-reported and identified according to the British National Formulary (age 44/45)^10^

Not at all/not very happy (vs very/fairly happy) (age 33)

Question: “How you feel	about life so far: all things considered, how happy are you?

*The response scale was reversed in the analysis, so the higher score represents worse wellbeing.

References corresponding to different measures:

1.	Rutter M, Tizard J, Whitmore K. Education, Health and Behaviour. London: Longmans; 1970.

8.	Bὂrsch-Supan A, Brugiavini J, H., , Makenbach J, Siegrist J, Weber G. Health, ageing and retirement in Europe. First results from the survey of health, ageing and retirement in Europe (SHARE). Manheim Research Institute for the Economics of Ageing (MEA);2005.

9.	Wiggins RD, Brown M, Ploubidis GB. A measurement evaluation of a six item measure of quality of life (CASP6) across different modes of data collection in the 1958 National Child Development Survey (NCDS) Age 55 years. Working paper 2017/2. Centre for Longitudinal Studies;2017.

10.	Joint Formulary Committee. BNF 68: September 2014-March 2015. London: Pharmaceutical Press; 2014.

| eTable 3. Correlations between continuous (r) and binary (rho) outcomes collected at the same age. | | | | | | | | | | | | | | | |
| --- | --- | --- | --- | --- | --- | --- | --- | --- | --- | --- | --- | --- | --- | --- | --- |
| **Continuous outcomes (r)** | | | | | | | | | **Binary outcomes (rho)** | | | | | | |
| **Age 23** | | | | | | | | | | | | | | | |
|  | | Malaise Inventory | | | |  | | |  | Poor/fair health | Malaise Inventory≥4 | | | | |
| Malaise Inventory | | 0.16 | | | |  | | | Seen MH specialist | 0.37 | 0.48 | | | | |
|  | |  | | | |  | | | Poor/fair health |  | 0.53 | | | | |
| **Age 33** | | | | | | | | | | | | | | | |
|  | | Malaise Inventory | | | |  | | |  | Poor/fair health | Malaise Inventory≥4 | | Not at all/not very happy | | |
| Malaise Inventory | | 0.34 | | | |  | | | Seen MH specialist | 0.33 | 0.51 | | 0.29 | | |
|  | |  | | | |  | | | Poor/fair health |  | 0.45 | | 0.23 | | |
|  | |  | | | |  | | | Malaise Inventory≥4 |  |  | | 0.52 | | |
| **Age 42** | | | | | | | | | | | | | | | |
|  | | Life satisfaction | | GHQ-12 | | | | |  | Poor/fair health | Malaise Inventory≥4 | | GHQ≥4 | | |
| Malaise Inventory | | 0.37 | | 0.56 | | | | | Seen MH specialist | 0.32 | 0.54 | | 0.45 | | |
| Life satisfaction | |  | | 0.34 | | | | | Poor/fair health |  | 0.45 | | 0.35 | | |
|  | |  | | | | | | | Malaise Inventory≥4 |  |  | | 0.66 | | |
| **Age 44/45** | | | | | | | | | | | | | | | |
|  | | | CIS-R - Anxiety | | | | | |  | CIS-R - Anxiety≥2 | Psychotropic medications | | | | |
| CIS-R - Depression | | | 0.33 | | | |  | | CIS-R - Depression≥2 | 0.64 | 0.17 | | | | |
|  | | |  | | | |  | | CIS-R - Anxiety≥2 |  | 0.16 | | | | |
| **Age 50** | | | | | | | | | | | | | | | |
|  | Malaise Inventory | | | | Life satisfaction | | CASP | WEMWBS |  | Poor/fair health | | Malaise Inventory≥4 | | SF-36≤52 | WEMWBS≤40 |
| SF-36 | 0.68 | | | | 0.52 | | 0.67 | 0.71 | Seen MH specialist | 0.45 | | 0.62 | | 0.61 | 0.43 |
| Malaise Inventory |  | | | | 0.42 | | 0.54 | 0.50 | Poor/fair health |  | | 0.52 | | 0.49 | 0.45 |
| Life satisfaction |  | | | |  | | 0.59 | 0.49 | Malaise Inventory≥4 |  | |  | | 0.77 | 0.62 |
| CASP |  | | | |  | |  | 0.69 | SF-36≤52 |  | |  | |  | 0.80 |
| *Note.* GHQ-12=General Health Questionnaire-12; CIS-R – Depression=Clinical Interview Schedule-Revised – Depression; CIS-R – Anxiety=Clinical Interview Schedule-Revised – Anxiety; SF-36=36-Item Short Form Survey; CASP=Quality of Life Scale; WEMWBS=Warwick-Edinburgh Mental Wellbeing Scale; Seen MH specialist=Seen mental health specialist.  The estimates are based on standardised outcomes and complete case analysis for each pair of measures. | | | | | | | | | | | | | | | |
